# Genetic and non-genetic components of family history of stroke and heart disease: a population-based study among adopted and non-adopted individuals

**DOI:** 10.1101/2023.05.28.23290649

**Authors:** Ernst Mayerhofer, Livia Parodi, Kaavya Narasimhalu, Andreas Harloff, Marios K Georgakis, Jonathan Rosand, Christopher D Anderson

## Abstract

**Background:** It is increasingly clear that genetic and non-genetic factors account for the association of family history with disease risk in offspring. We sought to distinguish the genetic and non-genetic contributions of family history of stroke and heart disease on incident events by examining adopted and non-adopted individuals.

**Methods:** We examined associations between family history of stroke and heart disease with incident stroke and myocardial infarction (MI) in 495,640 participants of the UK Biobank (mean age 56.5 years, 55% female) stratified by early childhood adoption status into adoptees (n=5,747) and non-adoptees (n=489,893). We estimated hazard ratios (HRs) per affected nuclear family member, and for polygenic risk scores (PRS) for stroke and MI in Cox models adjusted for baseline age and sex.

**Results:** 12,518 strokes and 23,923 MIs occurred over a 13-year follow-up. In non-adoptees, family history of stroke and heart disease were associated with increased stroke and MI risk, with the strongest association of family history of stroke for incident stroke (HR 1.16 [1.12, 1.19]) and family history of heart disease for incident MI (HR 1.48 [1.45, 1.50]). In adoptees, family history of stroke associated with incident stroke (HR 1.41 [1.06, 1.86]), but family history of heart disease did not associate with incident MI (p>0.5). PRS showed strong disease-specific associations in adoptees and non-adoptees. In non-adoptees, the stroke PRS mediated 6% risk between family history of stroke and incident stroke, and the MI PRS mediated 13% risk between family history of heart disease and MI.

**Conclusions:** Family history of stroke and heart disease increase risk for their respective conditions. Family history of stroke contains a substantial proportion of potentially modifiable non-genetic risk, indicating a need for further research to elucidate these elements for novel prevention strategies, whereas family history of heart disease represents predominantly genetic risk.

Graphic abstract

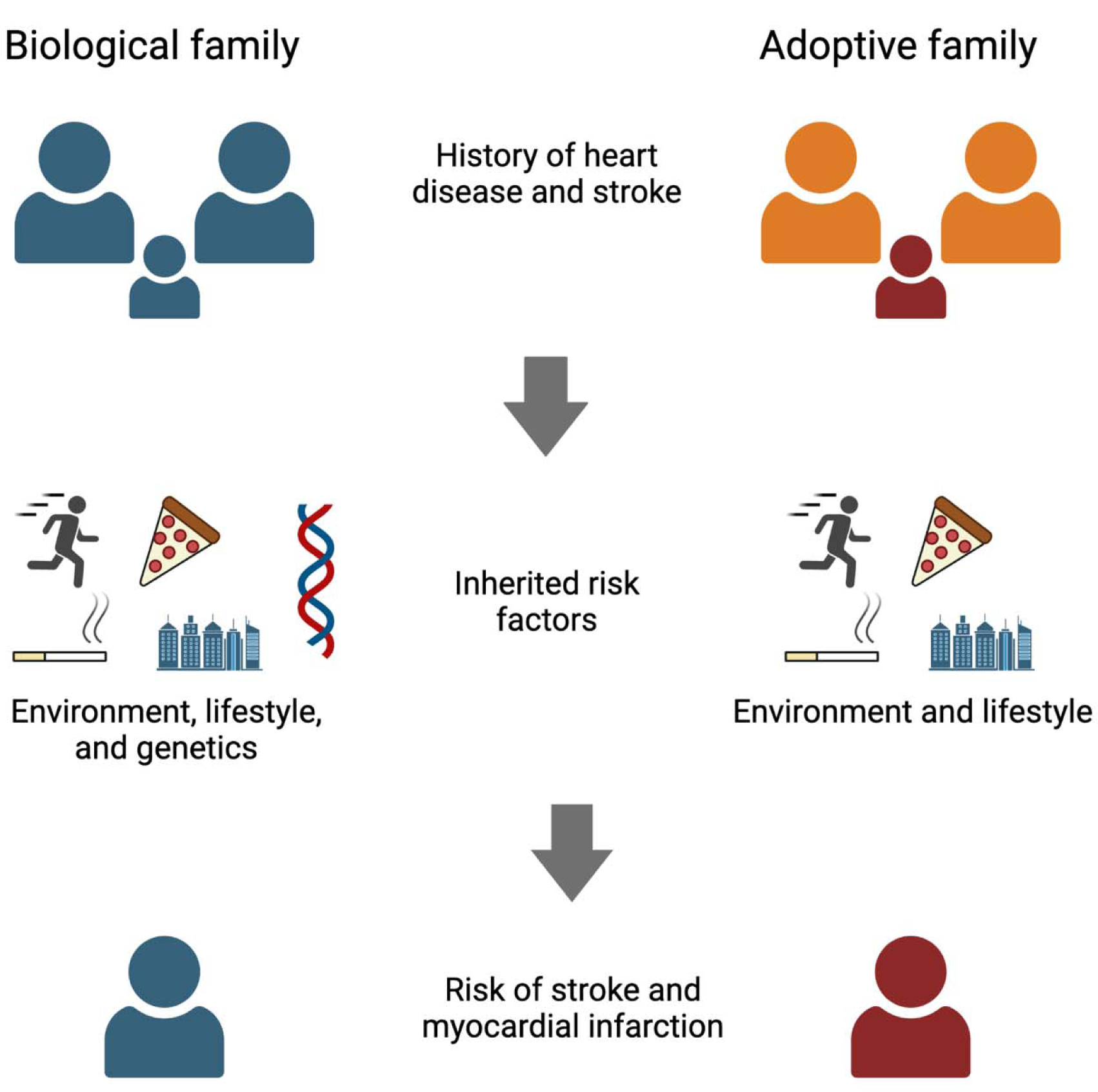

## Introduction

Stroke and coronary artery disease (CAD) are the leading causes of morbidity and mortality worldwide with rising incidence.^1,2^ Environmental, lifestyle, and genetic factors contribute substantially to their risk, and primary prevention efforts aim to control modifiable risk factors to decrease the burden of disease.^3^

The observation from epidemiological^4,5^ and twin studies^6,7^ that family history of stroke and CAD predispose individuals to a higher risk of developing these diseases has led to large international collaborations to study the genetic risk factors for stroke and CAD.^8,9^ Both stroke and CAD have been found to be highly polygenic, with heritability estimates, representing the variance explained by common variant genetics, of approximately ∼40% for stroke and ∼60% for coronary artery disease.^10,11^ Over time, genome-wide association studies (GWAS) have become large enough to identify common genetic variants that causally contribute to individual genetic risk of these common diseases. Polygenic risk scores (PRS) have been developed as a tool to capture the cumulative effect of multiple genetic variants on the risk of complex diseases and have gained association strength similar to traditional cardiovascular risk factors.^12,13^ As such, PRS have been investigated as potential clinical tools for risk prediction and stratification.^14^ Unlike family history, which represents the cumulative effects of the developmental family environment as well as the genetics of each parent and blood relative, genetic markers are constant throughout the lifespan and specific to the individual.

Previous studies support this assertion that family history and genetic risk may not be interchangeable terms.^15-17^ Depending on the heritability of the disease, family history might contain a substantial proportion of inherited non-genetic risk, such as smoking, dietary patterns, or general risk factor awareness, making it a partially modifiable risk factor. PRSs for many diseases mediate only a minor proportion of family history, and the association of PRS and family history on many health outcomes is largely independent.^16,18^ While such combined analyses of family history and PRSs suggest that family history contains a substantial proportion of non-genetic risk, they are limited by the performance of the PRS, which relies on the discovery power of the underlying GWAS and other challenges of PRS, such as portability,^19^ sensitivity to population stratification,^20^ and miscalibration.^21^

Individuals adopted early in childhood provide a unique opportunity to investigate the non-genetic risk contribution of family history, as they share lifestyle and environmental exposures with their adoptive families but not genetics. The objective of this study was to investigate whether family history for cardiovascular disease-related traits are specific for incident stroke and MI and how much of their risk is attributable to genetics. By examining the associations between family history, PRS, and the risk of stroke and MI in both biological and adopted families in a large cohort of individuals from the UK Biobank (UKB), we aimed to gain insight into the modifiability of disease-specific inherited risk, which could support efforts to engage in aggressive risk factor management and behavioral interventions in those with strong family history of disease.

## Methods

### Study population

The UKB is a population-based prospective cohort study that recruited over 500,000 participants aged 40-70 years between 2006 and 2010 from across the United Kingdom. Extensive baseline data, including sociodemographic, lifestyle, and health-related information were collected through questionnaires and physical measurements. Additionally, biological samples were obtained from each participant for genomic analyses. Genotyping was performed on two very similar platforms and imputation was performed on the Haplotype Reference Consortium for 488,377 participants.^22^ For the current study, we stratified individuals by adoption status (as defined by field 1767 gathered from the touchscreen question “Were you adopted as a child?”). Information about adoption circumstances, such as age at adoption, was not collected from study participants.

The UKB was approved by the North West Multi-centre Research Ethics Committee, and all participants provided informed consent. We accessed the data following approval of an application by the UKB Ethics and Governance Council (Application No. 36993).

### Family history

Family history was assessed through self-reported information from study participants at baseline. Illnesses of parents and siblings were gathered from study participants through multiple choice touchscreen questions. For individuals who indicated they were adopted, specific questions were employed to gather information regarding illnesses of adoptive parents and siblings (fields 20112-20114). In contrast, for non-adopted participants, the illnesses of biological parents and siblings were collected (fields 20107, 20110, and 20111). We gathered for each individual the number of adopted or biological nuclear family members with stroke and heart disease. We also gathered the number of adopted or biological parents (omitting siblings) with stroke and heart disease.

### Construction of genetic scores

We gathered summary statistics for stroke and MI from the most recent published GWAS that did not include data from UKB participants: from MEGASTROKE^23^ for stroke and from CARDIoGRAMplusC4D^24^ for MI. All GWAS were performed in individuals of European ancestry. We generated PRSs for those traits using PRS-CS, an unsupervised method which utilizes a high-dimensional Bayesian regression framework to derive a PRS from GWAS summary statistics without requiring an external validation cohort.^25^ This method outperforms traditional PRS approaches via its utilization of external linkage disequilibrium reference panels.^25^ PRS-CS with default parameters generated 1,108,218 SNP weights for stroke and 1,106,964 SNP weights for MI.

### Outcome assessment

The primary outcomes of interest were incident stroke and MI events. These outcomes were ascertained through linkage to national hospital admission and mortality registries. Stroke events were identified using International Classification of Diseases, 10th Revision (ICD-10) codes I60-I64, while MI events were identified using ICD-10 codes I21-I22, aligned with the diagnostic algorithm in the UKB (https://biobank.ndph.ox.ac.uk/showcase/ukb/docs/alg_outcome_main.pdf). Stroke subtypes are not available in the UK Biobank. Events that occurred after baseline were defined as incident events.

### Statistical methods and software

We used t-test, ANOVA, and Chi-Square test for comparison of continuous and discrete baseline variables, respectively. We used Cox proportional hazards models to estimate hazard ratios (HRs) and 95% confidence intervals (CIs) for the associations between family history and genetic scores and the risk of developing stroke and MI. In total, we used 4 exposure variables: the number of family members affected by heart disease and stroke, and the PRSs for stroke and MI.

Separate models were constructed with each exposure adjusted for age and sex, stratified by adoption status to explore potential differences in the associations. We also investigated potential effect modification by age and sex by including interaction terms in the models and conducting stratified analyses by age group and sex. We conducted mediation analyses to calculate the fraction of family history risk that was mediated by the PRSs. To maximize power, we performed the association models for family history in all individuals regardless of available genotyping data. Because the GWAS for stroke and MI were performed in European ancestry individuals, association models for the PRSs were restricted to individuals with available genetic data and of European ancestry. To rule out that the associations between family history and the outcomes were driven by those with unavailable genetic data or non-European ancestry, we performed sensitivity analyses for all models in those with available genetic data and European genetic ancestry to confirm robustness of our findings. We performed additional sensitivity analyses adjusted for cardiovascular risk factors at baseline: presence of hypertension, hyperlipidemia, diabetes, and smoking status (current/former/never smoker), and for family histories of stroke and heart disease of parents (without siblings) only.

SNP extraction and genetic score calculation was performed with PLINK,^26^ PRS-CS,^25^ and bcftools,^27^ relationship inference was performed with KING.^28^ All statistical analyses were conducted using R software, and a two-sided p-value < 0.05 was considered statistically significant. Mediation analyses were performed with the R package mediator (https://gerkelab.github.io/mediator/) that uses a counterfactual framework.^29^ Data extraction, curation, preparation, statistical analysis, and figure generation was done with RStudio.^30^

### Data availability

UKB participant data can be accessed by submitting an approved research proposal to UKB.^31^ The GWAS summary statistics used to create genetic scores are publicly accessible.^23,24^

## Results

### Study population characteristics

A total of 495,640 individuals were included in the analysis; 5,747 adoptees and 489,893 non-adoptees (**Figure 1**). The baseline characteristics of the study population are shown in **Table 1**. Adoptees were more likely to be current smokers and take lipid-lowering medications, had higher BMI, and had a lower prevalence of self-reported family history of stroke and heart disease; other statistically significant, but not clinically meaningful differences in HbA1c and the PRS were found. We found sex-specific differences in self-reported family history: both adopted and non-adopted women were more likely to report a positive family history for heart disease than men, non-adopted women were also more likely to report a family history of stroke than non-adopted men (**Table S1**). Over a median 13.4 years follow-up, 12,518 strokes and 23,923 MI events occurred. Adoptees were at higher absolute risk for an MI event (absolute risk 5.7% vs 4.8%, p=0.003).

**Figure 1.**
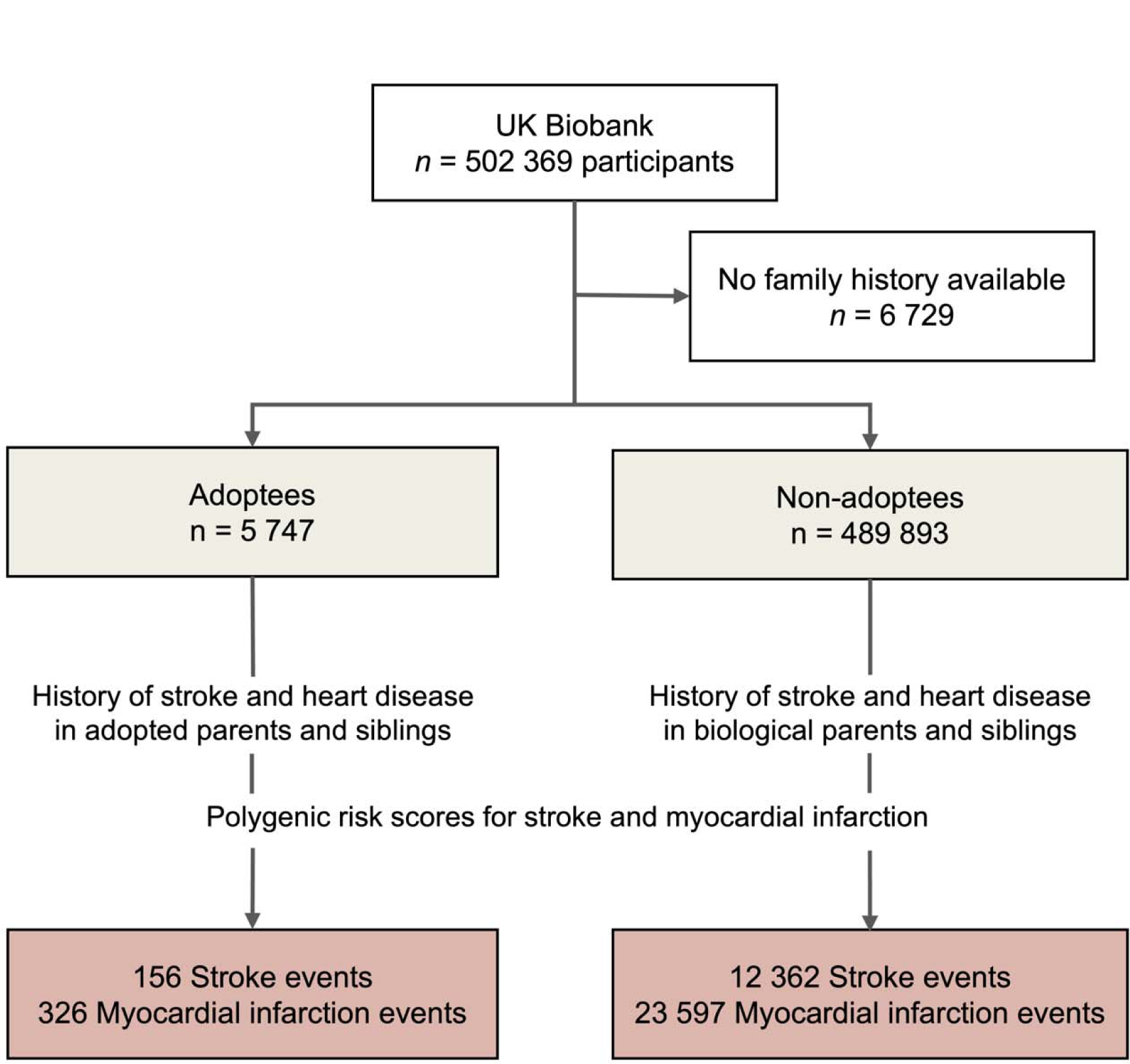
Study overview. Participants from the UK Biobank were stratified based on their self-identified adoption status during childhood. The association between family history of stroke and heart disease and polygenic risk scores for stroke and myocardial infarction with the incidence of stroke and myocardial infarction over a 13-year follow-up period were evaluated.

**Table 1:** Baseline characteristics by adoption status.

|  | Non-adoptees<br>(N=489,893) | Adoptees<br>(N=5,747) | p |
| --- | --- | --- | --- |
| Age at baseline, years, mean (SD) | 56.5 (8.08) | 56.0 (8.60) | <0.001 |
| Female sex, n (%) | 267,550 (54.6%) | 3,089 (53.7%) | 0.195 |
| White race, n (%) | 433,127 (88.4%) | 4,830 (84.0%) | <0.001 |
| European genetic ancestry, n (%) | 401,378 (81.9%) | 4,156 (72.3%) | <0.001 |
| Smoking status, n (%) |  |  | <0.001 |
| Never Smoker | 268,229 (54.8%) | 2,610 (45.4%) |  |
| Ex-Smoker | 168,864 (34.5%) | 2,148 (37.4%) |  |
| Current Smoker | 50,971 (10.4%) | 961 (16.7%) |  |
| Systolic blood pressure, mmHg, mean (SD) | 138 (18.6) | 138 (18.8) | 0.0442 |
| Body mass index, kg/m<sup>2</sup>, mean (SD) | 27.4 (4.80) | 28.1 (5.20) | <0.001 |
| Use of lipid-lowering medication, n (%) | 80,275 (16.4%) | 998 (17.4%) | 0.0482 |
| HbA<sub>1c</sub>, mmol/l, mean (SD) | 36.1 (6.73) | 36.7 (7.73) | <0.001 |
| Family history of stroke, n (%) | 136,946 (28.0%) | 1,346 (23.4%) | <0.001 |
| Family history of heart disease, n (%) | 221,991 (45.3%) | 2,245 (39.1%) | <0.001 |
| PRS for myocardial infarction, mean (SD) | -0.441 (0.125) | -0.435 (0.124) | 0.0029 |
| PRS for stroke, mean (SD) | -0.736 (0.0723) | -0.731 (0.0721) | <0.001 |
| Stroke events, n (%) | 12,362 (2.5%) | 156 (2.7%) | 0.381 |
| Myocardial infarction events, n (%) | 23,597 (4.8%) | 326 (5.7%) | 0.0029 |

HbA<sub>1c</sub>, Glycated hemoglobin, SD, standard deviation; PRS, polygenic risk score

### Stroke risk in biological and adopted families

In non-adoptees, both family history of stroke and heart disease were significantly associated with stroke risk, with a much stronger association for family history of stroke than for family history of heart disease (HR 1.16 [1.12, 1.19] vs 1.08 [1.06, 1.11] per one affected family member, **Figure 2A**). A one standard deviation (SD) increase in the stroke PRS increased the risk for stroke by an HR of 1.19 [1.17, 1.21]. Family history of stroke was significantly associated with the stroke PRS (0.052 [0.046, 0.058] SD increase in the stroke PRS per family member with a stroke).

**Figure 2.**
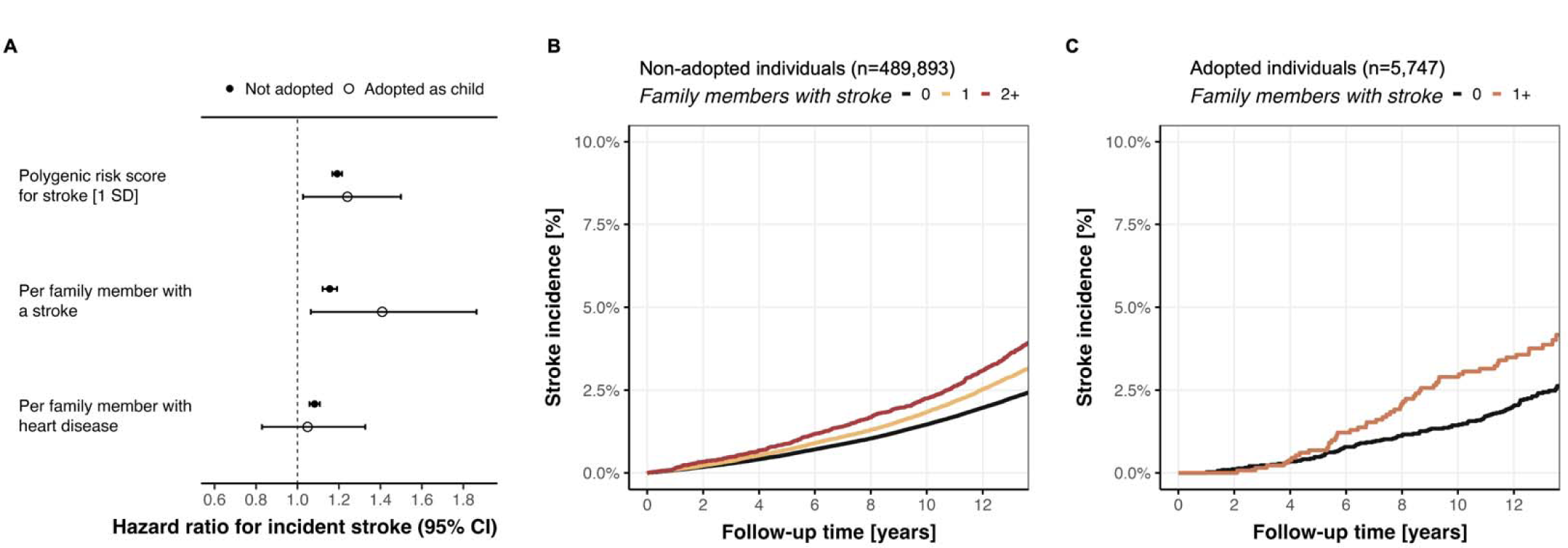
Comparison of association between family history of stroke and incident stroke among non-adoptees and adoptees. Family history for stroke increased risk for incident stroke both among non-adoptees and adoptees (A). Absolute incidence curves for stroke in non-adoptees (B) and adoptees (C) across strata of family members affected by a stroke. In adoptees, groups were collapsed because of the low number of people with two or more family members with a stroke. Family history was self-reported at baseline for biological and adopted father, mother, and siblings, respectively.

In the cohort of 5,747 adoptees, positive family history of stroke yielded an association with incident stroke even greater than non-adoptees, with an HR of 1.41 [1.06, 1.86] per affected family member. The stroke PRS had a similar association with stroke risk as in non-adoptees with a hazard ratio of 1.24 [1.03, 1.50]. When comparing absolute risk, we found a clear separation of the incidence curves in both non-adoptees (**Figure 2B**) and adoptees (**Figure 2C**). We found no association between family history of stroke and the stroke PRS in adoptees (p=0.49).

We found a stronger association between family history of stroke and incident stroke among younger non-adoptees, with a similar yet statistically insignificant trend observed in adoptees (**Figures S1-S2**). There was a stronger association of family history of heart disease with incident stroke in non-adopted women (p_interaction_=0.0037, **Figure S3**), but no sex-specific effects were found in adoptees (all p_interaction_>0.27, **Figure S4**). The PRS mediated 6% of the association between family history of stroke on incident stroke in non-adoptees, and 0% in adoptees. Sensitivity analyses confirmed the findings in fully adjusted models for cardiovascular risk factors, in the subgroup of individuals with available genetic data and European ancestry, and in the models that only considered the family history of the parents (**Figures S5-S7**).

### MI risk in biological and adopted families

In the cohort of 489,893 non-adoptees, both family history of heart disease and stroke were significantly associated with incident MI risk, with a much stronger association for family history of heart disease than for stroke (HR 1.48 [1.45, 1.50] vs HR 1.05 [1.03, 1.08] per one affected family member, **Figure 3A**). A one SD increase in the MI PRS increased the risk for MI by an HR of 1.43 [1.41, 1.46]. Family history of heart disease was significantly associated with the MI PRS (0.119 [0.115, 0.124] SD increase in the MI PRS per family member with heart disease).

**Figure 3.**
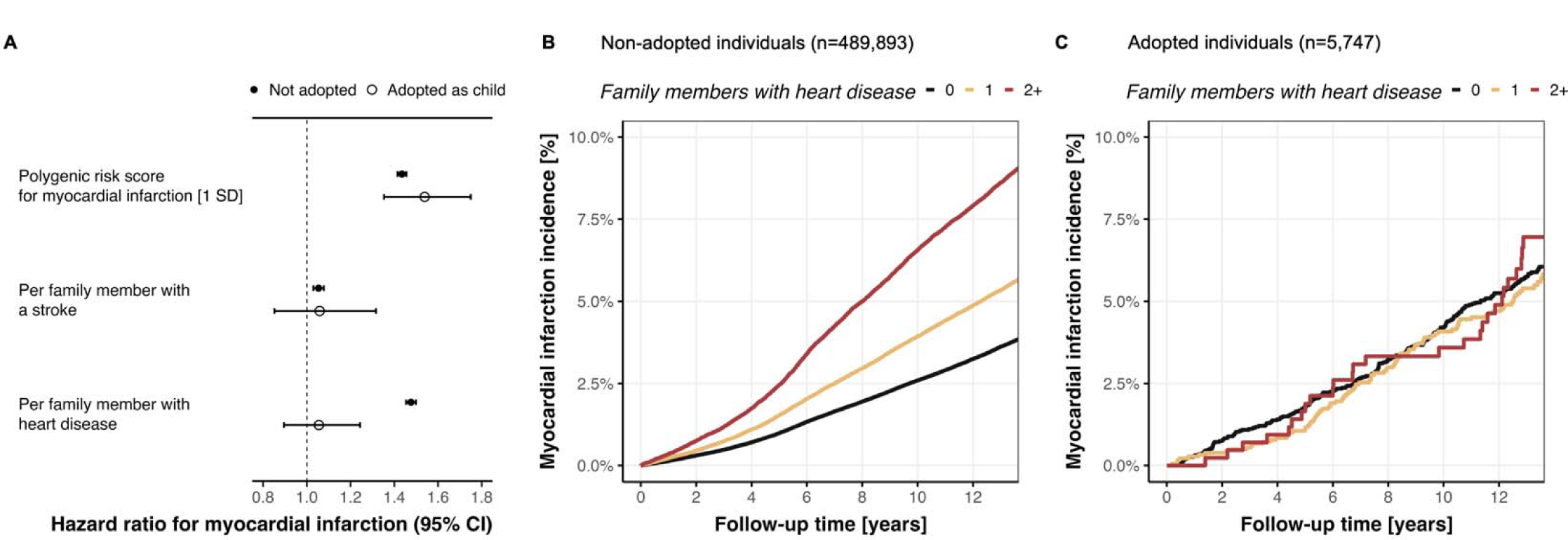
Comparison of association between family history of illnesses related to cardiovascular disease and incident myocardial infarction among non-adoptees and adoptees. Family history for cardiovascular-related traits was only associated with incident stroke among non-adoptees, not among adoptees (A). Absolute incidence curves for stroke in non-adoptees (B) and adoptees (C) across strata of family members affected by heart disease. Family history was self-reported at baseline for biological and adopted father, mother, and siblings, respectively.

In the cohort of 5,747 adoptees, neither positive family history of heart disease nor stroke was associated with MI (p=0.52 and p=0.61, respectively, **Figure 3A**). The MI PRS had a similar association with MI risk as in non-adoptees with a hazard ratio of 1.54 [1.35, 1.75]. When comparing absolute risk across strata of family history of heart disease, we found a clear separation of the incidence curves in non-adoptees (**Figure 3B**), but not in adoptees (**Figure 3C**). We found no association between family history of heart disease and the MI PRS in adoptees (p=0.65).

We found stronger effect sizes for family history for stroke and heart disease and the MI PRS in younger non-adoptees (all p_interaction_<0.001, **Figure S8**), but no age-dependent association in adoptees (**Figure S9**). Sex-specific analyses yielded a better MI PRS performance in non-adopted men (p_interaction_=0.021, **Figure S10**), but no interactions with sex in adoptees (all p_interaction_ >0.09, **Figure S11**). The MI PRS mediated 13% of the association between family history of heart disease on incident MI in non-adoptees, and 0% in adoptees.

Sensitivity analyses confirmed the findings in fully adjusted models for cardiovascular risk factors in the subgroup of individuals with available genetic data and of European ancestry, and in the models that only considered the family history of the parents (**Figures S12-S14**).

## Discussion

In this study, we investigated the association between family history of stroke and heart disease, PRS for each, and the incidence of stroke and MI in the UKB among individuals with biological and adopted parents. We found that self-reported family history for stroke and heart disease are disease-specific risk factors for incident stroke and MI in biological families. Family history of stroke was also a strong risk factor for incident stroke among adoptees, suggesting that it contains a substantial proportion of non-genetic risk. In contrast, family history of heart disease followed primarily genetic risk, as evidenced by a much larger mediation by the MI PRS and no association with MI risk in adoptees, despite greater statistical power for MI outcomes. Both family history and genetic risk showed a stronger association among younger individuals, suggesting that inherited risk factors might indicate risk earlier in life.

The disease-specific associations between family history of stroke and CAD is potentially due to the diverse etiologies of stroke compared to the relatively more homogeneous pathogenesis of CAD. Stroke has multiple etiologies, each with distinct risk factors and underlying pathophysiology;^32,33^ in contrast, CAD is primarily driven by atherosclerosis.^34^ Although risk factors for CAD and stroke overlap, they are different in magnitude.^35^ These differences could explain the variation in genetic and environmental contributions observed in our study when comparing family history of stroke and heart disease. Furthermore, the diversity in stroke subtypes may also account for the stronger disease-specific association observed for CAD family history compared to stroke family history, as the latter may encompass a wider range of causal factors.

Although family history of stroke is associated with genetic risk for stroke among biological families, it contains a substantial proportion of non-genetic and thus potentially modifiable risk, as it increases the risk for stroke in adoptees by a similar magnitude as in biological offspring. No such observation was observed for CAD, confirming results from previous studies with similar design.^36,37^ Our results carry practical implications, as they indicate that family history of stroke is an indicator of a higher burden of modifiable risk factors rather than predominantly genetic risk, offering insights for primary prevention strategies. Inquiring about family history is a rapid, straightforward approach to stratify individuals for targeted primary prevention interventions. Our results show that depending on the answer to a few simple questions, absolute risk differs up to three-fold over the subsequent 13 years, with different magnitudes for stroke and MI. Based on our data, this information could serve as a foundation for determining which patients may benefit from more aggressive risk factor management. Instead of conveying a genetic predisposition with no actionable outcomes, family history of stroke could function as a motivating factor, fostering awareness and encouraging lifestyle modifications or even more aggressive risk factor targets in primary prevention.

In our study, we observed that adoptees had a higher burden of cardiovascular risk factors and related outcomes. There is evidence for a higher burden of behavioral and psychological health problems leading to elevated rates of alcohol, smoking, and substance abuse in adoptees^38^, as well as increased all-cause mortality in adoptees compared to biological offspring^39^ linked to age at adoption^40^. Because we stratified our analyses by adoption status, the overall elevated risk among adoptees should not have biased our results. Although we did not specifically focus on this aspect, our results highlight the importance of tailored risk factor management strategies targeting adoptees for prevention efforts to reduce their elevated risk and improve overall cardiovascular health.

Age-stratified analyses revealed that inherited risk factors might exert a stronger influence on stroke and MI risk in younger individuals. Since we used self-reported family history at inclusion, younger participants had presumably younger parents and thus a positive family history in those individuals may be indicative of a high burden of genetic or environmental risk factors. This observation implies that early identification of individuals with a family history of stroke or heart disease, particularly among younger populations, could facilitate more effective targeting of preventive interventions. In addition, we found that women were more likely to report family history than men. Previous studies have found similar sex-specific behavioral bias,^41,42^ indicating that family history questions should be tailored according to gender and might be more useful for risk stratification for women than men due to elevated response rates.

Our study has several limitations. First, the PRSs were derived from European populations, potentially limiting the generalizability of the associations between the PRSs and outcomes to other populations. However, the concept of PRS is not limited by genetic ancestry but rather by the available GWAS data. Furthermore, by using a PRS based on common genetic variants, we might have underestimated genetic risk for stroke and MI transmitted by other genetic and epigenetic mechanisms, such as rare variants,^43^ methylation patterns,^44^ genetic imprinting,^45^ among others. But since our primary focus was not the actual performance of the PRSs, but rather on using them as tools to compare family history and markers of inherited risk, this should not have biased our findings. Additionally, since we were unable to use the most up-to-date GWAS due to its overlap with UKB participants, our PRSs might have been underpowered compared to performance of most recent PRS and thus the mediation estimates of PRS on family history may have been underestimated.^8,46,47^ Second, our analysis relied on self-reported family history, introducing recall and reporting biases. Nonetheless, this reflects real-world conditions, and there is evidence that self-reported family history for cardiovascular-related traits is reliable compared to ascertainment through disease registries or relative’s self-report.^48,49^ Third, mediation estimation in Cox models remains an area of active development and may warrant further methodological investigations.^50^ Finally, although we adjusted for potential confounders, residual confounding remains a possibility.

## Conclusion

Our study demonstrates the importance of distinguishing between family history of stroke and heart disease in the context of incident stroke and MI risk assessment. Our findings further show that family history and genetic risk are not interchangeable terms as only a small part of family history is mediated by common variant genetic risk. They further demonstrate different subtypes of vascular risk carried by family history of stroke and MI, respectively, with different genetic and environmental contributions. Our study underscores the potential value of family history of stroke as a partially modifiable risk factor, and highlights the need for targeted primary prevention efforts, particularly in younger high-risk populations and adoptees. Although our study has limitations, it provides valuable insights into the complex interplay between genetic risk, family history, and the incidence of stroke and MI. Future studies should evaluate the feasibility and the benefit of intensified risk factor management in individuals with positive family history of stroke.

## Data Availability

UKB participant data can be accessed by submitting an approved research proposal to UKB. The GWAS summary statistics used to create genetic scores are publicly accessible.

## Acknowledgments

This research has been conducted using the UK Biobank Resource under Application Number 36993. Figure 1 was created with Biorender.com.

