## Supplemental Material for "Genetic and non-genetic components of family history of stroke and heart disease: a population-based study among adopted and non-adopted individuals"

| **Not adopted individuals** | **Female (N=267,550)** | **Male (N=222,343)** | **p** |
| --- | --- | --- | --- |
| Family history of stroke, n (%) | 77,319 (28.9%) | 59,627 (26.8%) | <0.001 |
| Family history of heart disease, n (%) | 127,121 (47.5%) | 94,870 (42.7%) | <0.001 |
| **Adopted individuals** | **Female (N=3,089)** | **Male (N=2,658)** | **p** |
| Family history of stroke, n (%) | 725 (23.5%) | 621 (23.4%) | 0.949 |
| Family history of heart disease, n (%) | 1,276 (41.3%) | 969 (36.5%) | <0.001 |

**Supplemental Table S1. Comparison of self-reported family history by sex, stratified by adoption status.**

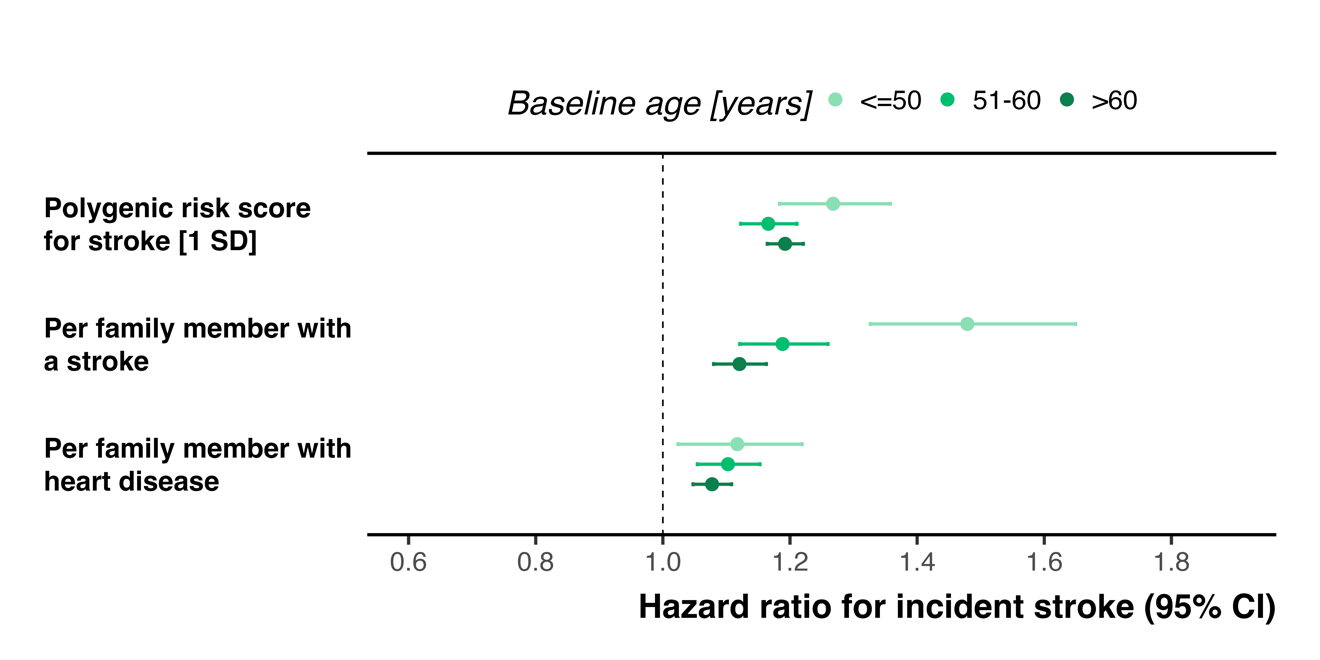

**Supplemental Figure S1.** Associations between self-reported family history illnesses and the stroke PRS with incident stroke stratified by age at baseline in non-adopted individuals.

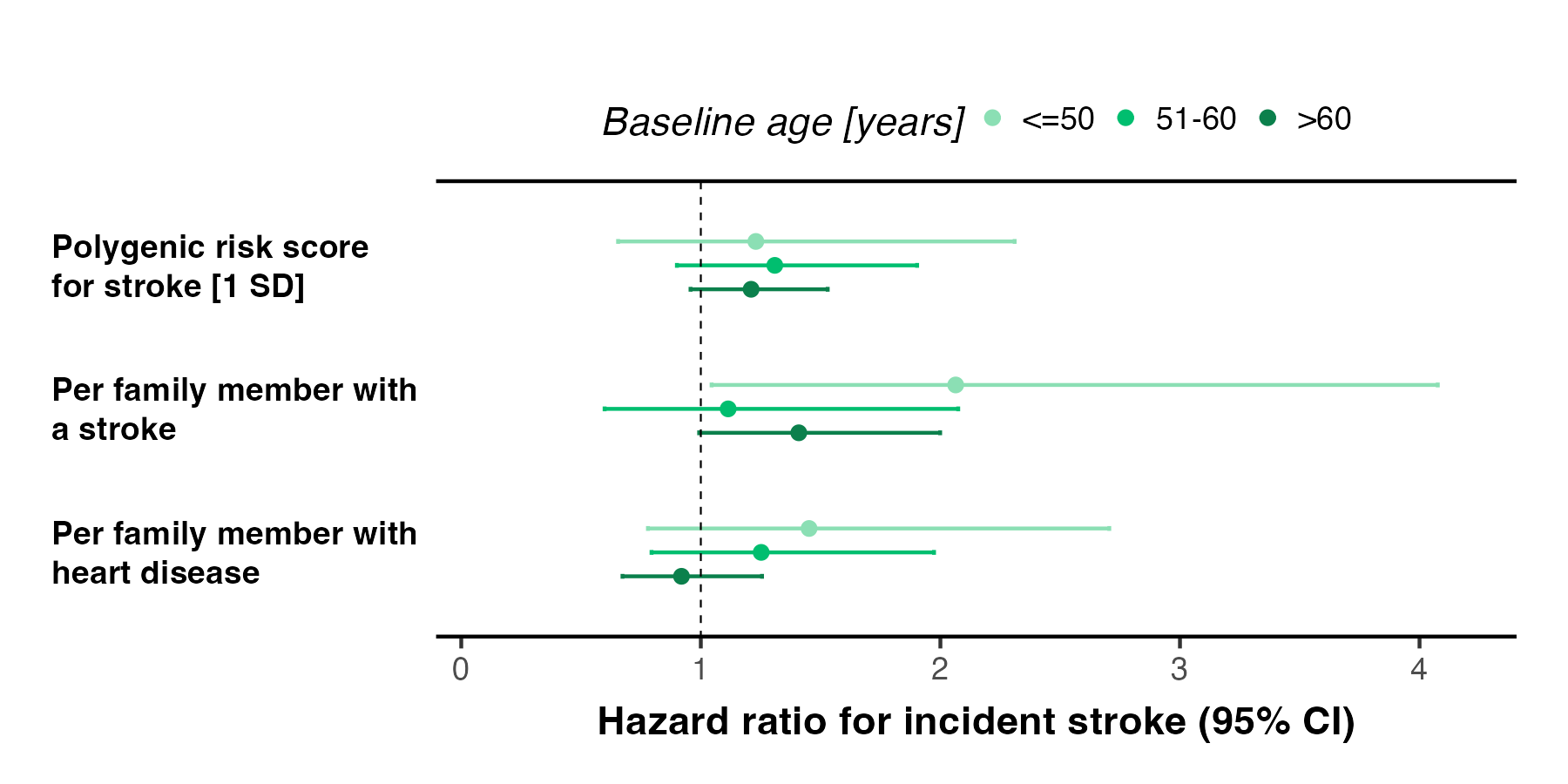

**Supplemental Figure S2.** Associations between self-reported family history illnesses of adopted family members and the stroke PRS with incident stroke stratified by age at baseline in adopted individuals.

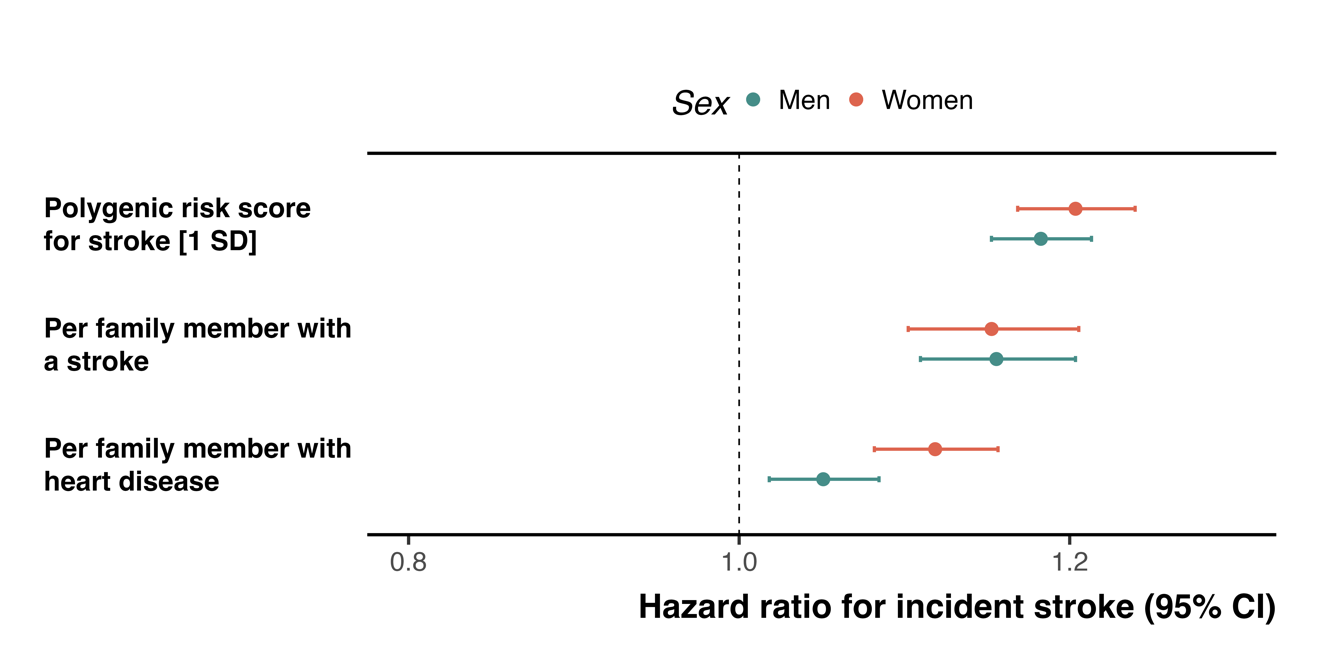

**Supplemental Figure S3.** Associations between self-reported family history illnesses and the stroke PRS with incident stroke stratified by sex in non-adopted individuals.

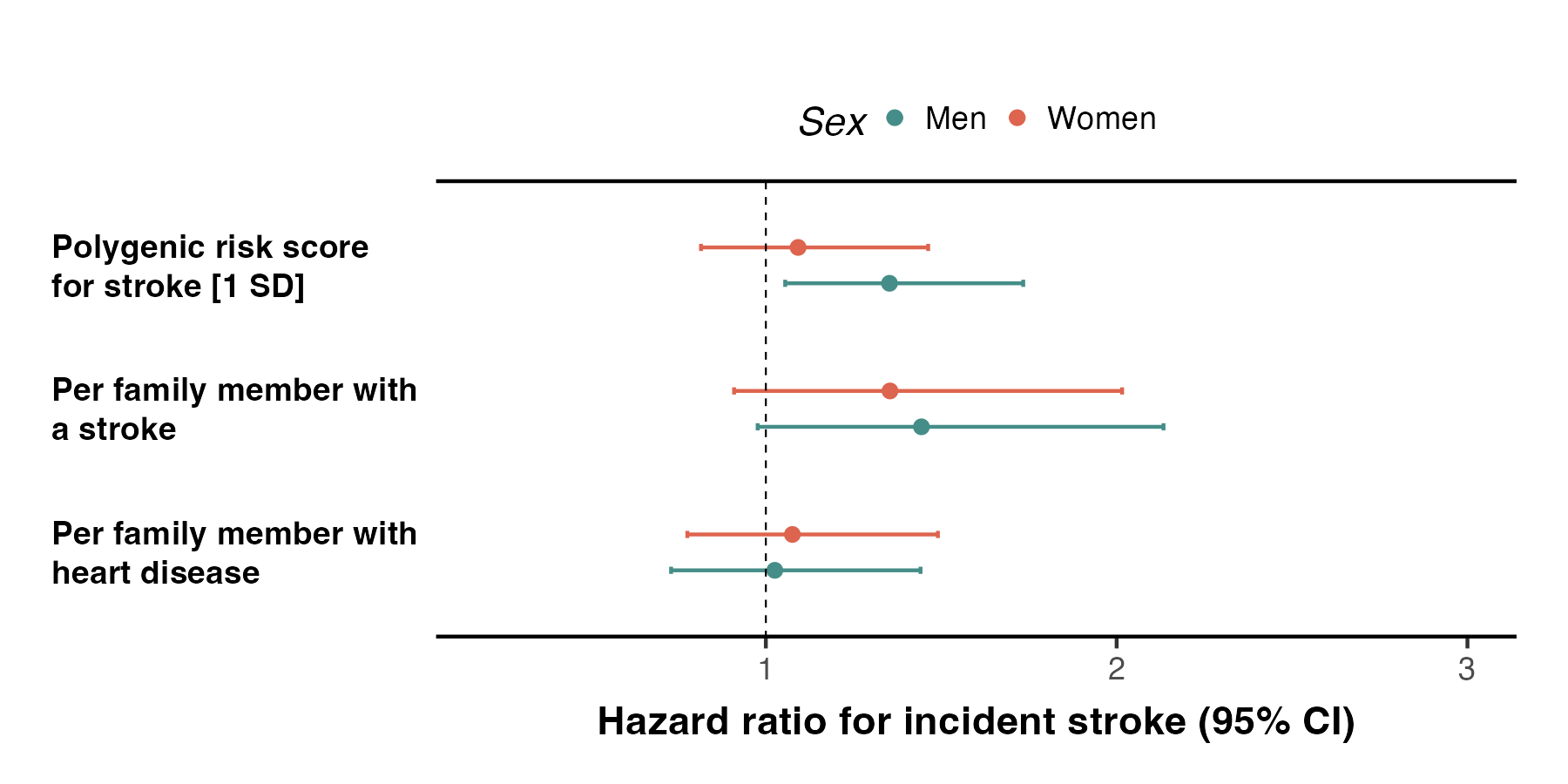

**Supplemental Figure S4**. Associations between self-reported family history illnesses of adopted family members and the stroke PRS with incident stroke stratified by sex in adopted individuals.

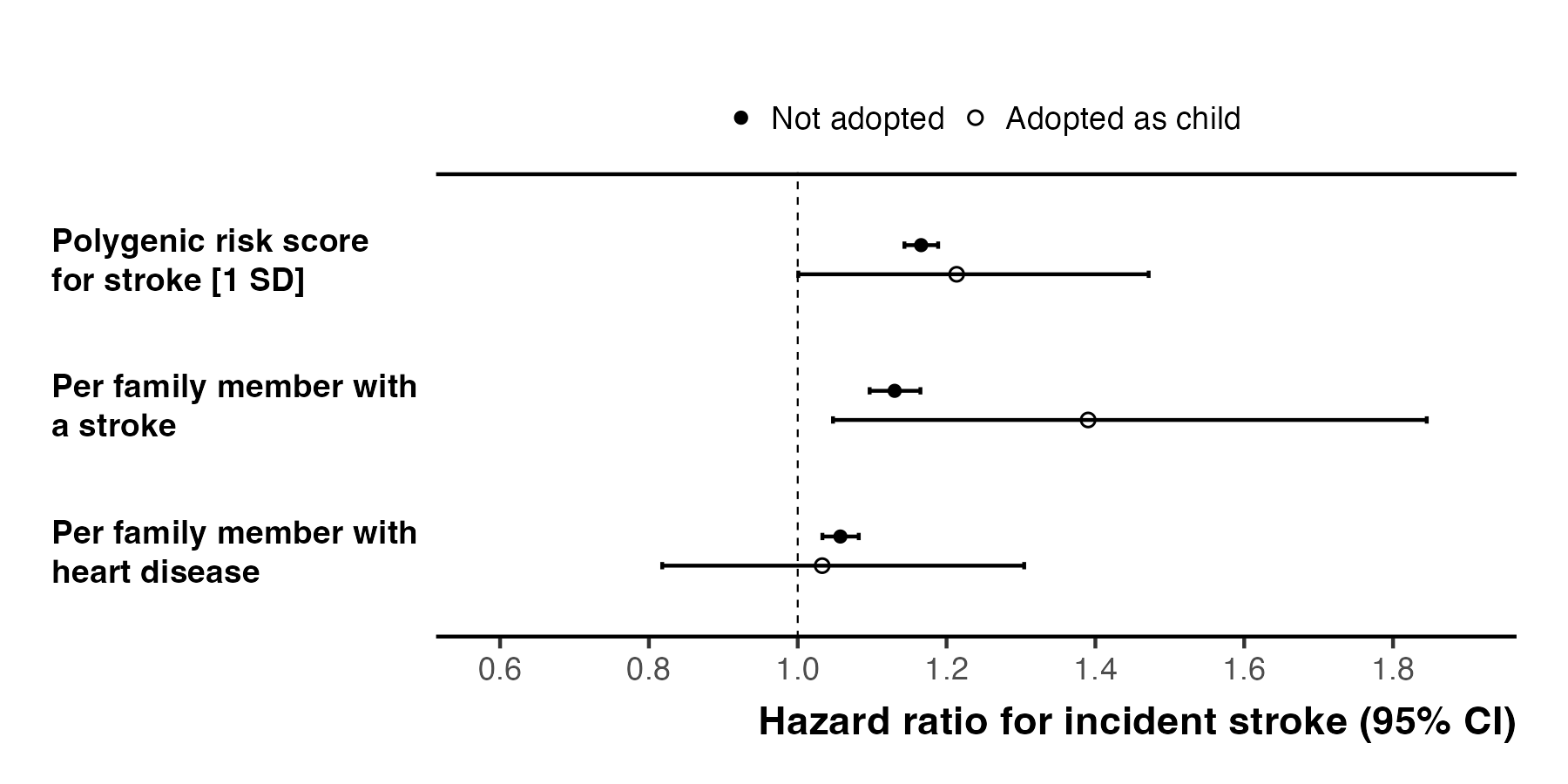

**Supplemental Figure S5.** Associations between self-reported family history illnesses and the stroke PRS with incident stroke in fully adjusted models for cardiovascular risk factors.

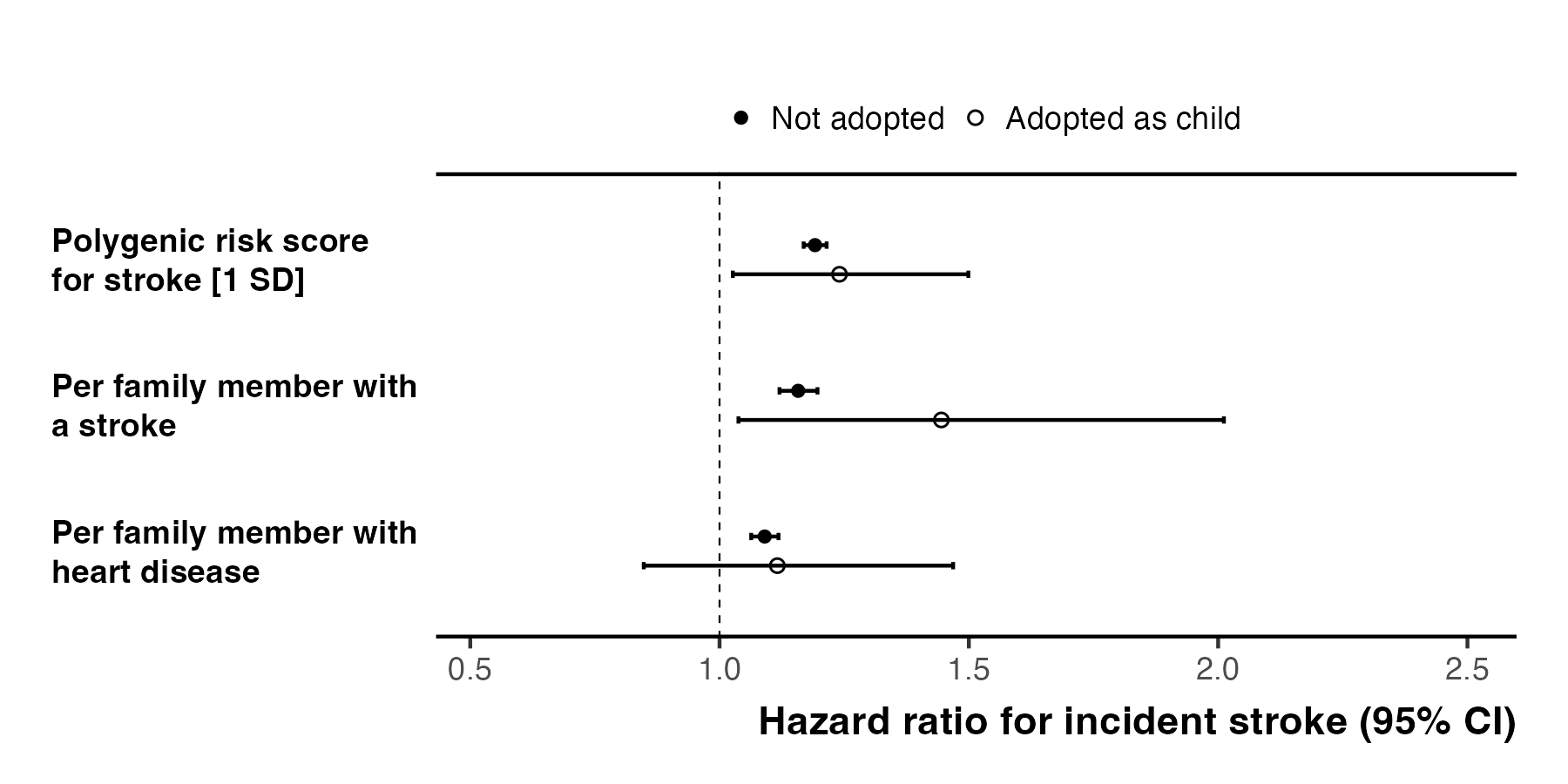

**Supplemental Figure S6.** Associations between self-reported family history illnesses and the stroke PRS with incident stroke in the subpopulation of people with available genetic data and of European genetic ancestry.

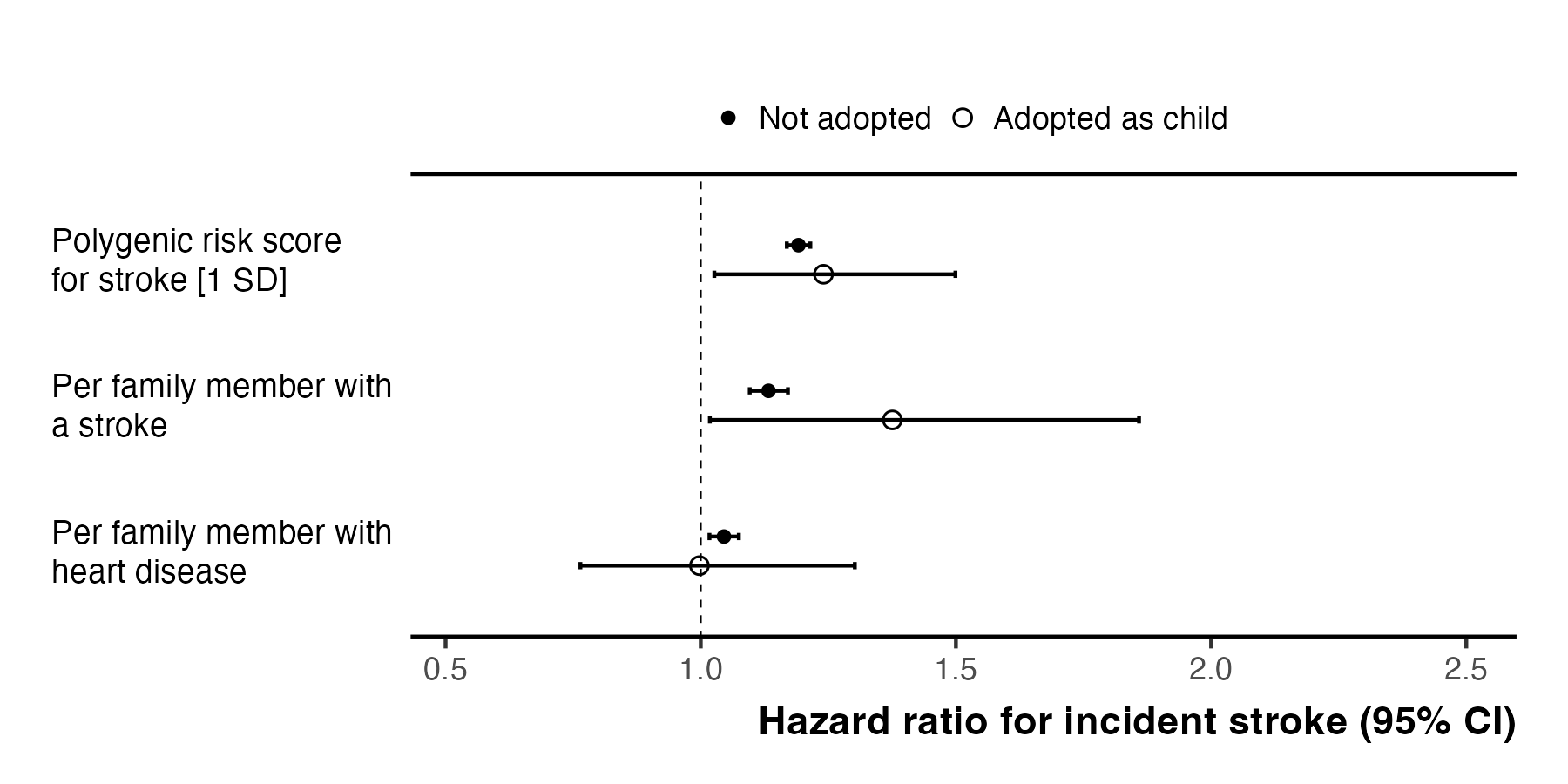

**Supplemental Figure S7.** Associations between self-reported family history illnesses of parents only (without considering siblings) and the stroke PRS with incident stroke.

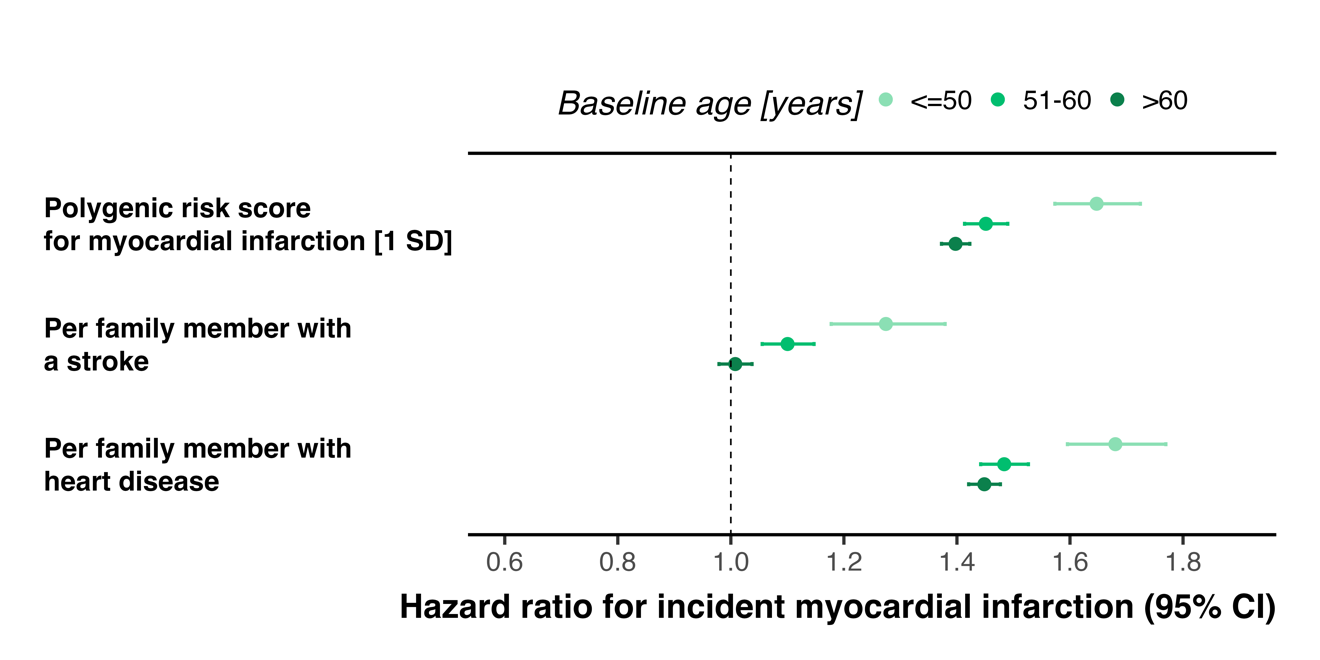

**Supplemental Figure S8.** Associations between self-reported family history illnesses and the MI PRS with incident MI stratified by age at baseline in non-adopted individuals.

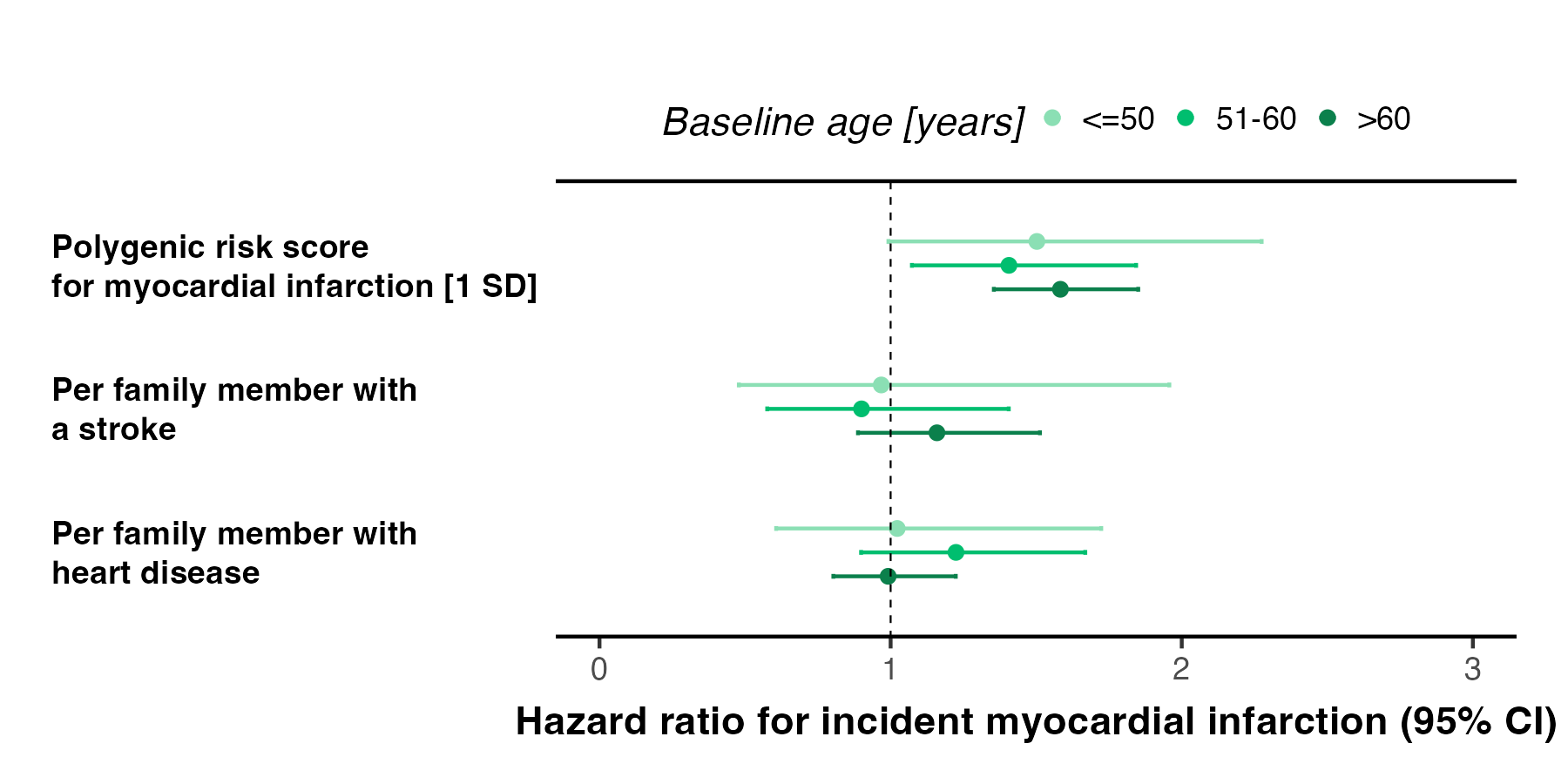

**Supplemental Figure S9.** Associations between self-reported family history illnesses and the MI PRS with incident MI stratified by age at baseline in adopted individuals.

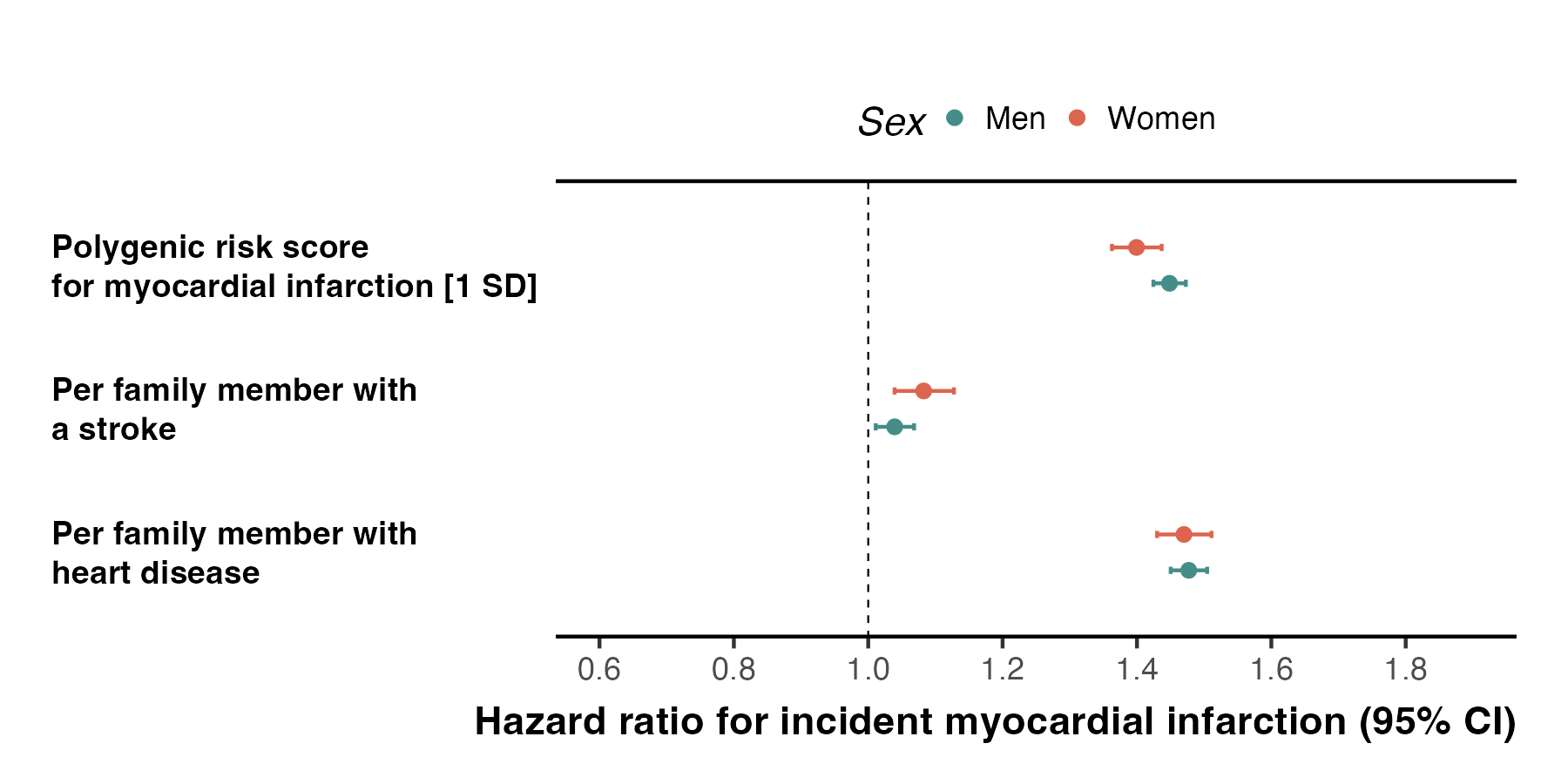

**Supplemental Figure S10.** Associations between self-reported family history illnesses and the MI PRS with incident MI stratified by sex in non-adopted individuals.

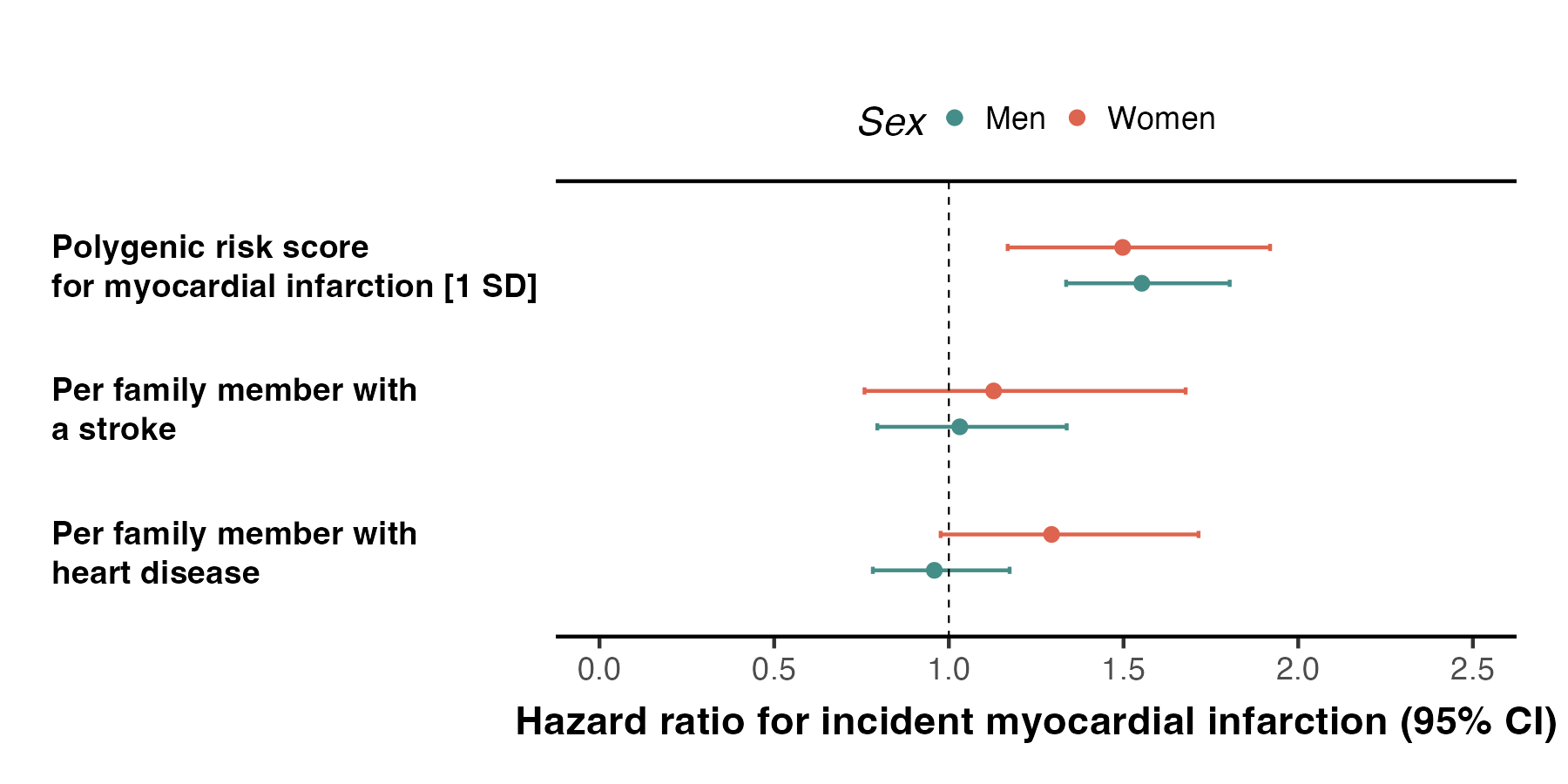

**Supplemental Figure S11.** Associations between self-reported family history illnesses and the MI PRS with incident MI stratified by sex in adopted individuals.

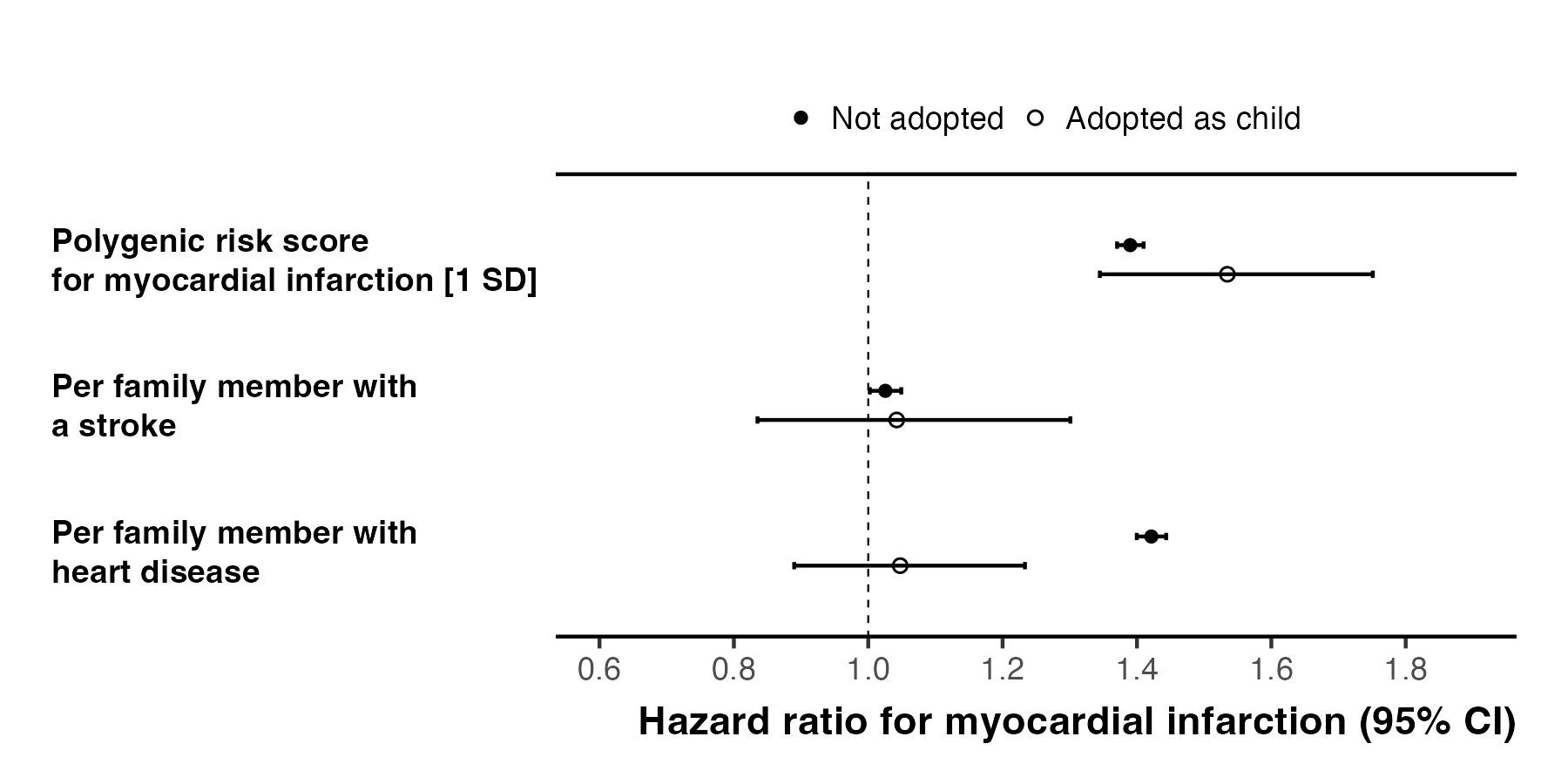

**Supplemental Figure S12.** Associations between self-reported family history illnesses and the MI PRS with incident MI in fully adjusted models for cardiovascular risk factors.

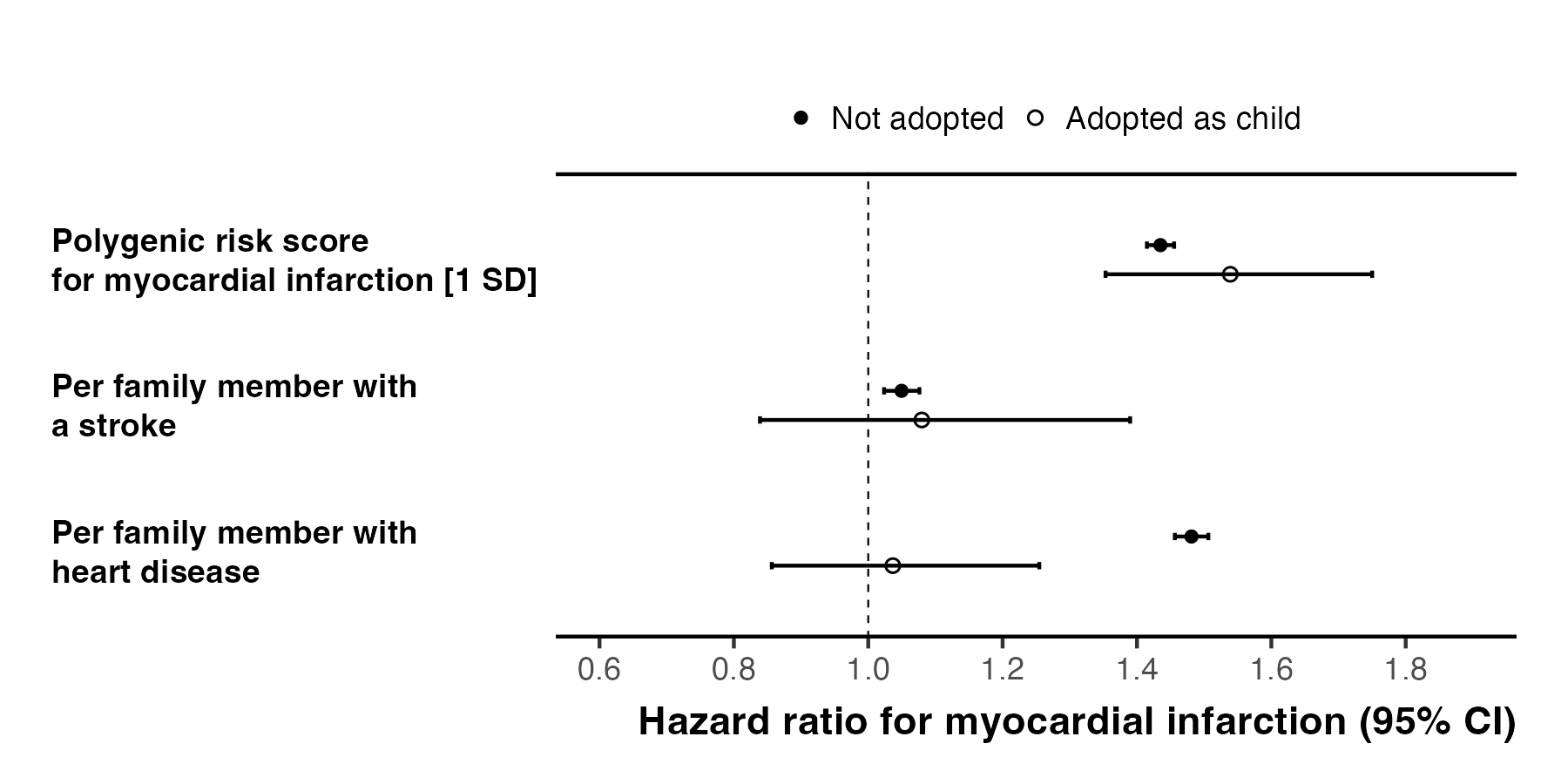

**Supplemental Figure S13.** Associations between self-reported family history illnesses and the MI PRS with incident MI in the subpopulation of people with available genetic data and of European genetic ancestry.

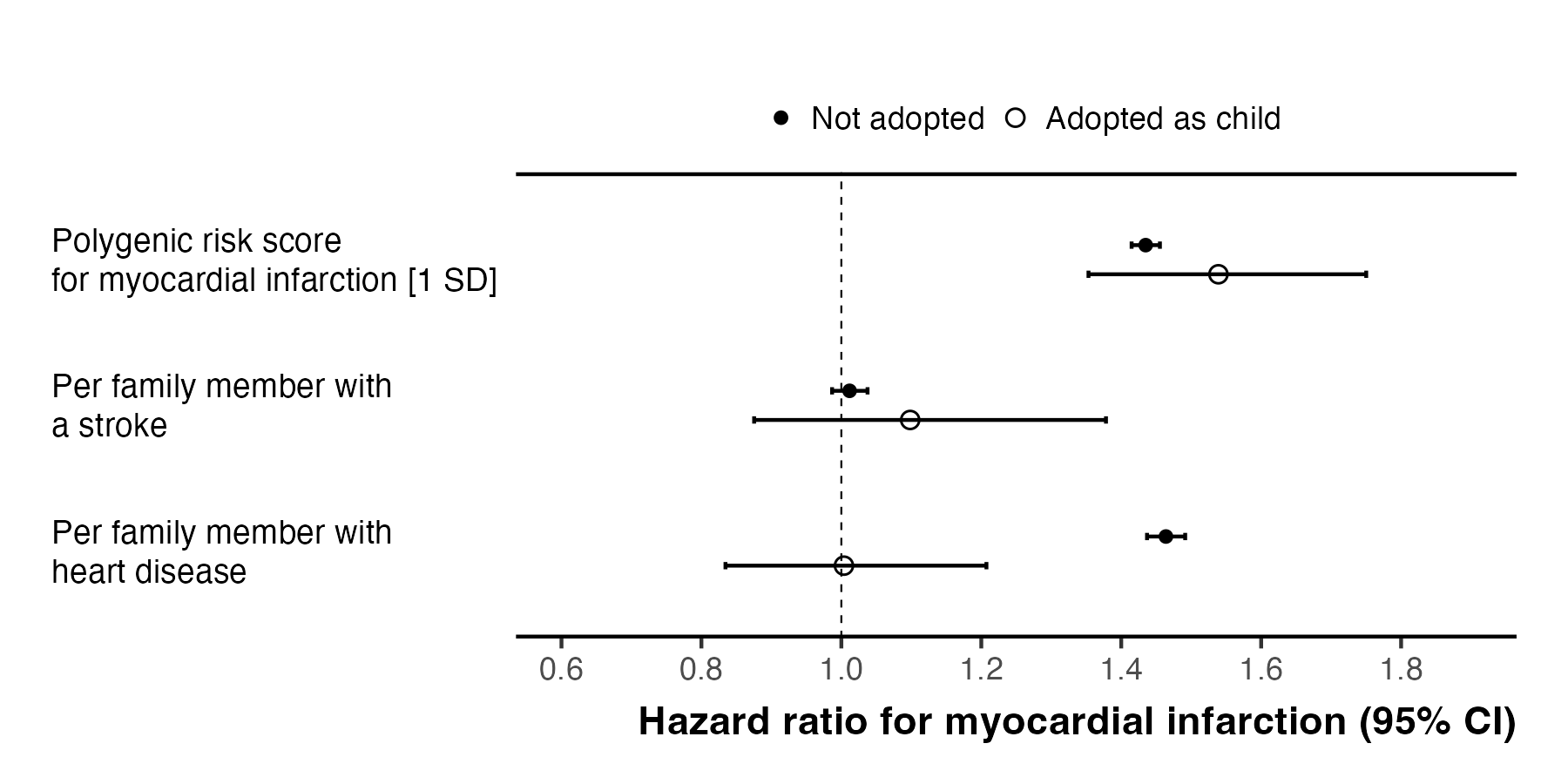

**Supplemental Figure S14.** Associations between self-reported family history illnesses of parents only (without considering siblings) and the MI PRS with incident MI.
